# A compositional approach to the analysis of white blood cell counts for early COVID-19 detection

**DOI:** 10.1101/2025.05.27.25328204

**Authors:** Zhilong Zhang, Jan Graffelman, Márcio Dorn

**Affiliations:** Department of Biostatistics, University of Washington; Department of Statistics and Operations Research, Universitat Politècnica de Catalunya; Institute of Informatics, Center of Biotechnology, Federal University of Rio Grande do Sul

**Keywords:** log-ratio transformation, compositional data, classification model, principal component analysis, linear discriminant analysis, biplot, ROC curve, AUC

## Abstract

The investigation of using complete blood-count (CBC) data for COVID-19 infection diagnosis has been a topic of interest in the last couple of years. It could be used as an affordable complementary tool to RT-qPCR and is particularly useful for developing areas struggling to test their population for suffering from massive COVID-19 infection. However, previous research on using CBC data for COVID-19 infection classification didn’t appreciate the compositional nature of white blood cell counts data. In this paper, we treat white blood cell counts data as compositional variables and apply compositional data visualization methods, using biplots based on log-ratio principal component analysis. Also, we apply compositional classification models to detect COVID-19 infection, using log-ratio linear discriminant analysis. We successfully illustrate the efficacy of compositional methods by building a compositional classification model superior to traditional models and highlight the benefits of analyzing CBC data from a compositional perspective. In a database of symptomatic individuals, we achieve a classification rate of 85% for the PCR-test result using the main CBC composition with some additional blood characteristics.

## 1 Introduction

The public health crisis of Coronavirus disease (COVID-19) is one of the most severe pandemics humans have experienced for the past few centuries [10], and it still causes large burdens to our society in many ways including economy, health services, life expectancy, etc. [11]. Nowadays, although we have RT-qPCR as the gold standard for diagnosing SARS-Cov2 infection [3], it remains impossible to employ this test massively in certain developing areas due to financial and administration problems. There exist other available diagnostic tests, such as antigen tests, which may be more accessible in areas with limited resources. On the other hand, in most scenarios those developing areas like Ecuador and Brazil need a well-functioning surveillance system more than others to avoid massive infection and control health care expenditure.

Motivated by this situation, we are trying to use the complete blood-count (CBC) data analysis as a cheap and efficient complementary tool to help us diagnosing potential COVID-19 infected patients [4]. In this classification problem, we have all of the common blood test results/indicators as our predictors and COVID-19 infection status (binary, given by RT-qPCR) as our outcome. The first step is to apply principal component analysis (PCA) [8] to the whole CBC dataset and the five major white blood cells (WBC) counts for preliminary data visualization and possible feature selection. Based on the prior knowledge, we know that the five major types of white blood cell counts (see Figure 1 for the five major WBC counts: neutrophils, eosinophils, basophils, monocytes, and lymphocytes) will give us high diagnostic/prediction power for many diseases as they are closely connected with immune response [15].

**Figure 1:**
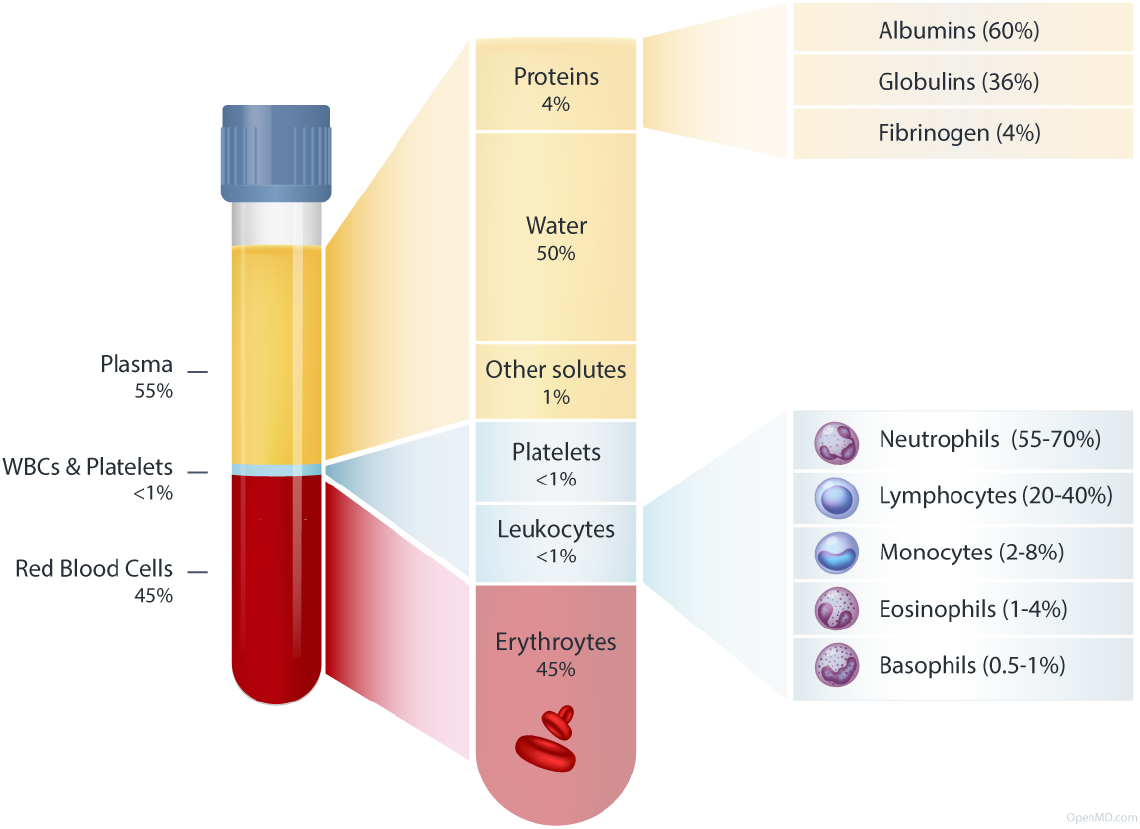
The components of blood (source: https://openmd.com/guide/blood-components).

Compositional data is a special type of data that is affected by a linear constraint which complicates the classical analysis, and which can be dealt with by applying the log-ratio transformation[1]. Importantly, compositional data analysis is ratio-based and focuses on the relative information in the counts, contrary to standard methods that are based on differences. Classical multivariate methods like PCA and linear discriminant analysis (LDA) have been adapted to the compositional context [2][7][5]. The five major types of white blood cell counts can be viewed as natural compositional data as each of the five WBC counts is part of a whole, the total WBC count. LDA can be applied to the raw WBC dataset and the log-ratio transformed WBC compositional data. By doing so, we are able to compare the performance of LDA on the raw data and the compositional data and discuss the advantage of combining compositional data analysis and traditional classification method like LDA in this COVID-19 detection scenario.

The remainder of this article is organized as follows. We first give some background on compositional data analysis with a review of log-ratio principal component analysis and log-ratio discriminant analysis, followed by the analysis of an empirical CBC database[13] for Ecuador residents. This article finishes with a discussion and some recommendations for the analysis of compositional datasets like the WBC dataset.

## 2 Methodology

In this section, we briefly introduce the statistical methods used in this paper, including compositional data and the log-ratio transform, log-ratio principal component analysis (LR-PCA), and log-ratio discriminant analysis (LR-LDA).

### 2.1 Compositional Data

Compositional data consists of variables that are parts of some whole. Proportions, percentages, and concentrations are common types of compositional data. When analysing those type of datasets, some issues come up that are hard to overcome by traditional statistical methods. For example, it is hard to define correlation and independence with respect to compositional data because the data is subject to a linear constraint. This motivates us to build new methods and techniques to deal with this specific type of data, generally called methods for compositional data analysis (CoDA) [16].

To be more specific, let us define a *composition* of *D* parts to be a random row vector:

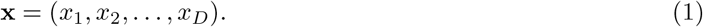

And we define the domain of our compositional data **x** to be a *simplex S*^*D*^:

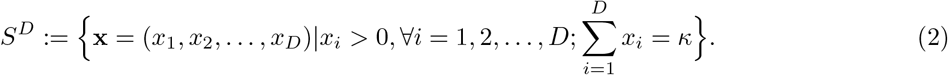

Since we only care about the relative values of compositional data, we can re-express our data to a relative constant *κ*_0_ (normally *κ*_0_ = 1), and this operation is called *closure* operation 𝒞 :

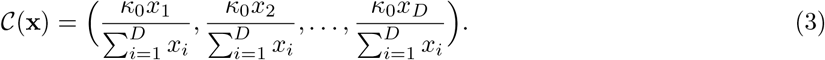

On the domain (*S*^*D*^) where our compositional data lives, we need new a geometry instead of the Euclidean geometry in ℝ^*n*^ due to the lack of linear vector space structure and corresponding metric. To equip *S*^*D*^ with a new vector space structure and the corresponding new geometry (called Aitchison geometry [1]), we define “addition” and “multiplication” in *S*^*D*^, called perturbation and powering respectively:

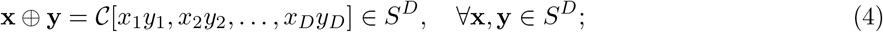

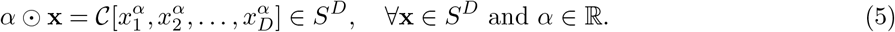

It’s easy to see that *S*^*D*^ with those two operations is an abelian group with an external product, and therefore a vector space. To measure distance and angle on *S*^*D*^, we define the Aitchison inner product:

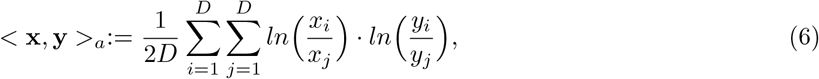

and we can define norm and distance induced by it.

Due to the nature of compositional data, three principles are proposed and should be fulfilled by any statistical method that we are trying to apply to compositional data [16]. They are *scale-invariance* (multiplying a composition by any positive value, or equivalently selecting any positive constant *κ*_0_ shouldn’t matter for the results of our analysis), *permutation invariance* (reordering parts of the composition **x** should give us equivalent results in compositional data analysis), and *subcompositional coherence* (any analysis applied to a subcomposition should give us results coherent with the analysis applied to the full composition. For example, a well defined distance should satisfy that the distance between two sub-compositions is always less or equal than the distance between the two corresponding full compositions).

Compositional data satisfy the three principles by first applying the *log-ratio transformation* to the compositional data and then apply classical statistical method to the transformed data. Three common log-ratio transformation are the additive log-ratio transformations [16] (*alr*):

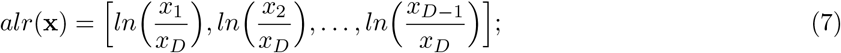

the centred log-ratio transformation [16] (*clr*):

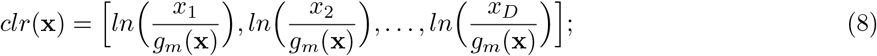

where *g*_*m*_(**x**) is the geometric mean of the components of the composition **x**:

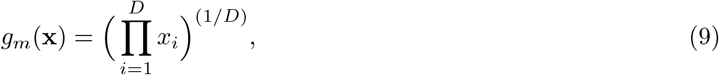

and the isometric log-ratio transformation [16] (*ilr*):

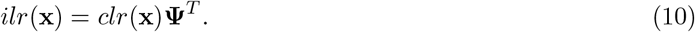

In the definition of *ilr*, **Ψ** is a contrast matrix with dimension (*D* − 1) × *D*. This matrix satisfies:

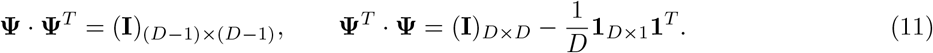

Notice that all three log-ratio transformations have their advantages and disadvantages. For example, *ilr* is an isometry mapping between *S*^*D*^ and ℝ^*D−*1^ but is computationally heavier, and its coordinates’ expressions are based on the choice of a basis (or equivalently, the choice of **Ψ**) thus they are not unique. Also, for *alr* one needs to specify the denominator in Equation (7) because our analysis should satisfy permutation-invariance thus the position of components doesn’t matter.

### 2.2 LR-PCA

We introduce Log-Ratio PCA (LR-PCA) as the method for exploring compositional data. The log-ratio transformation part is chosen to be *clr* transformation (Equation (8)) although we can combine PCA with other log-ratio transformations as well. The advantage of choosing *clr* includes the uniqueness of our transformation (there is no need to choose a contrast matrix **Ψ**, unlike *ilr* in Equation (10); or a denominator for *alr* in Equation (7)), preserving isometry between Aitchison distance of compositions and Euclidean distance of transformed coordinates (unlike *alr* in Equation (7)), etc. One thing to notice is that since every component in our composition naturally has the same unit, we prefer a covariance-based PCA after the log-ratio transformation to a correlation-based PCA.

To be more specific, let us present LR-PCA in a sample based manner. Therefore, for the mean, covariance and variance symbols presented below, we are referring to the sample mean, sample covariance and sample variance respectively. Also, unless otherwise specified, we define all vectors as column vectors to avoid notation confusion. Assume we have *n* observations (**x**_1_, **x**_2_, …, **x**_*n*_) that are sampled independently from the same distribution (i.i.d. samples), and each has dimension *D*. We denote the data matrix as **X**_*n×D*_. Continuing with notations introduced earlier, the coordinate of *clr*(**x**)_*i*_ equals:

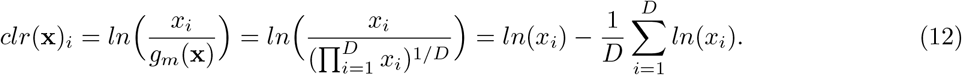

We note that the *clr* transformation amounts to a centering operation of the logarithmic scores. To write it in a matrix form, we have the log-transformed data **X**_*l*_:

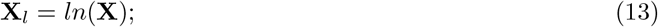

and the *clr* -transformed data **X**_*clr*_:

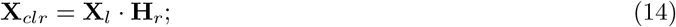

and the column centered *clr* data **X**_*cclr*_:

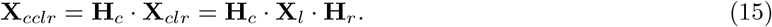

Where

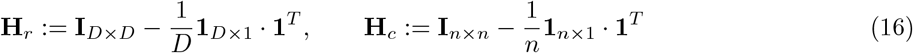

are the centering matrix in the *clr* definition and before the PCA process respectively, and we call our double-centered log-transformed data **X**_*cclr*_. After the double-centering process, we can just apply standard

PCA to **X**_*cclr*_, either by the spectral decomposition of its covariance matrix or by the SVD of **X**_*cclr*_.

Using the singular value decomposition, we can write

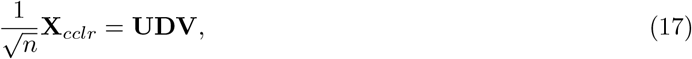

where **U** is the *n* × *r* matrix of left singular vectors (**U**^*T*^ **U** = *I*), **D** is the *r* × *r* diagonal matrix with non-increasing singular values, **V** is the *D* × *r* matrix of right singular vectors (**V**^*T*^ **V** = *I*) and *r* is the rank of **X**_*cclr*_.

Then we are able to construct form biplots based on

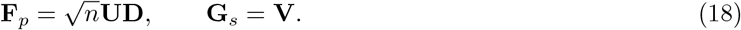

By using 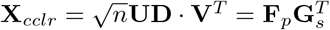, we are able to draw a biplot called *form biplot*. In this way, we plot **F**_*p*_ as row coordinates and **G**_*s*_ as the column coordinates (transformed coordinates of unit axis vector in **R**^*D*^). Selecting the first two columns of principal components **F**_*p*_ as data points and rows of the first two eigenvectors (which are the first two column of **G**_*s*_) as arrows enable us to draw a two-dimensional biplot. This plot captures most of the information limited to two dimensions to visualize this multivariate dataset. Moreover, the form biplot preserves the Euclidean distance in the following sense:

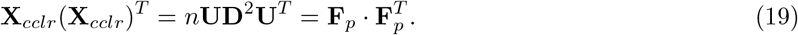

We can also define alternative biplot coordinates:

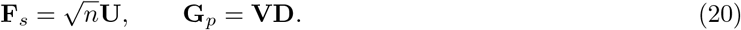

By using 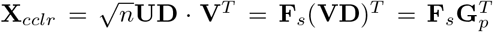, we are able to draw another biplot called the *covariance biplot*. In this way, we plot **F**_*s*_ as the row coordinates and **G**_*p*_ = **VD**_*s*_ as the column coordinates. One property of **G**_*p*_ is that it equals the covariance between variables and components:

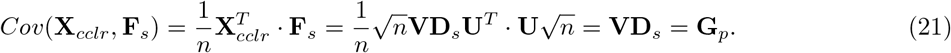

Therefore, selecting the first two columns of standardized principal components **F**_*s*_ as data points and the covariances of variables and the first two standardized components (which are the first two column of **G**_*p*_) as arrows enable us to draw another two-dimensional plot. Moreover, Euclidean distances in the covariance biplot represent Mahalanobis distances of the original observations in the following sense:

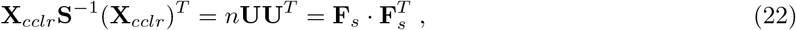

where **S** is the sample covariance 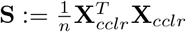.

There are some special properties of a compositional biplot obtained by LR-PCA [2] that deserve to be mentioned. First is the isometry of *clr* and the distance preserving property of the form biplot ensures that the Euclidean distance between LR-PCA scores equals the Aitchison distance between data points:

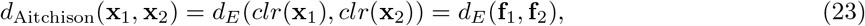

where **f**_1_, **f**_2_ are the two rows of **F**_*p*_. This equation can be easily checked using the definition of the Aitchison distance and the *clr* transformation.

Also, it is easy to see from the centering process that the origin in both biplots represents the geometric mean of the compositions. And the links between LR-PCA vectors represent pair-wise log-ratios: if *x*_1_ and *x*_2_ are the first and the second part of a compositional variable, then we have

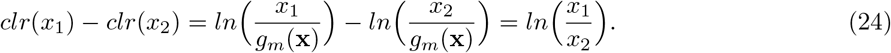

The length of a link between two vectors represents the standard deviation of the corresponding log-ratio:

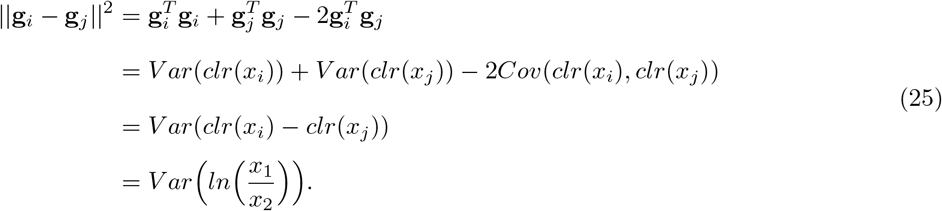

Based on Equation (24) and Equation (25), we have the following observations for a compositional biplot obtained by LR-PCA: close to coincident biplot vectors suggest proportionality of parts; cosines of angles between links represent correlations between log-ratios; and collinear biplot vectors suggest a one-dimensional pattern for a subcomposition.

### 2.3 LR-LDA

For a compositional dataset, we can not directly apply the LDA method for classification because of the structural singularity of our covariance matrix **Σ**. However, we can use a generalized inverse (e.g. the Moore–Penrose inverse) to overcome this problem ([17]). Also, in order to make our method applicable to multi-class cases instead of two group datasets, we introduce log-ratio linear discriminant analysis (LR-LDA) in the framework of Fisher’s multi-class discriminant analysis.

To be more specific, let us introduce Fisher’s multi-class discriminant analysis first. Fisher’s discriminant analysis aims to find a linear transformation **a** of our random vector **x** that maximizes the ratio of variability between groups to variability within groups.

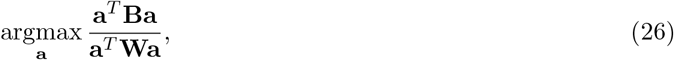

where **B** is defined as the variability between *k* groups and **W** is defined as the variability within *k* groups:

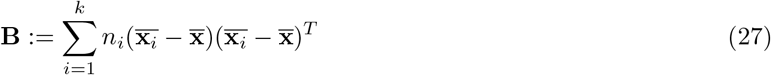

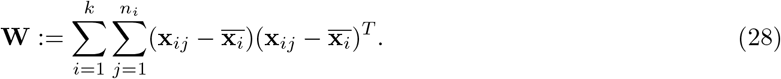

We denote *n*_*i*_ as the sample size of group *i* and *n* as the total sample size of whole dataset. Also, 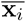 is the mean vector of group *i* and 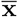 is the total mean vector of the whole dataset. **x**_*ij*_ is the data vector of case *j* that belongs to the group *i*. Notice that we have the following decomposition of the total variability, expressed by the total sum of squares:

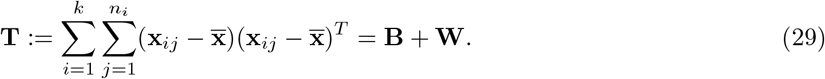

We choose to solve the objective function (26) by using the singular value decomposition. For the ease of notation, let us assume we have centered the dataset (this is automatically true after the double-centered log-ratio transformation as in Equation (15)). Define the diagonal matrix **D**_*w*_ := diag (*n*_1_*/n, n*_2_*/n*, …, *n*_*k*_*/n*) and 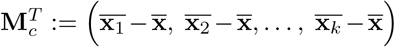. Then LDA can be performed by the singular value decomposition:

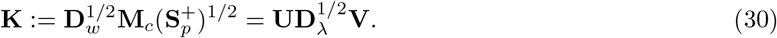

Notice that 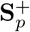 is the Moore-Penrose inverse of **S**_*p*_, the pooled covariance matrix, which is defined as a reweighed version of **W**:

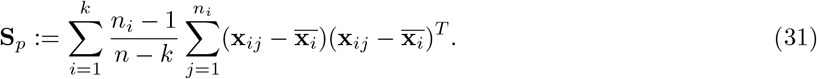

The SVD method is attractive since it provides a way to draw biplots, as now we have the row coordinates of the group means and the column coordinates of transformed vectors:

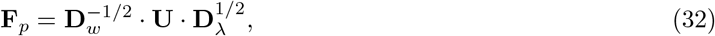

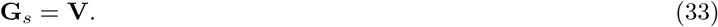

Now we can project our compositional dataset **X**_*cclr*_ introduced in Equation (15) to the optimal linear transformation and obtain the scores of the individual observations for LR-LDA classification in *min*(*D* − 1, *k* − 1) dimensions:

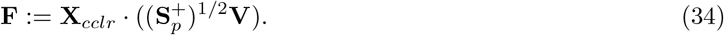

With the classification scores of our dataset, we can compare them with the group mean coordinates and decide their predicted group based on the Euclidean distance from the group mean in the discriminant space. To be more specific, we have the Euclidean distance between score of the individual observation **F**_*j*_ (*j*-th row of **F**, *j* = 1, 2, …, *n*) and the score of *i*-th group mean 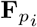 (*i*-th row of **F**_*p*_, *i* = 1, 2, …, *D*):

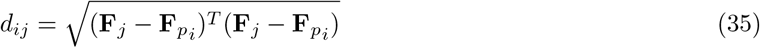

The predicted group of case *j* is the one that minimizes the distance over all *i*: argmin{*d*_1*j*_, *d*_2*j*_, …, *d*_*Dj*_}. Alternatively, classification can be based on the posterior probabilities, which are derived from the linear classifier assuming a normal distribution with the same covariance matrix but different means for the *k* groups.

## 3 Database and Data Cleaning

In this section, we introduce our dataset and its variables, with some summary statistics. Moreover, we clean our dataset to exclude abnormal medical cases.

### 3.1 Variable description

Our original CBC dataset was obtained between 2021/03/03 to 2021/08/09 in Ecuador, collected by Segurilab and Previne Salub laboratories [4]. It consists of 375 observations (clinical cases) with 24 variables, including demographic variables (sex, age), CBC results for each patient (including leukocytes counts, WBC counts and percentages, mcv, mch, etc.) and their infection status. Table 1 shows all variables in our dataset with brief descriptions.

**Table 1:** Variables with brief descriptions of the Ecuador CBC database.

| Variable names | Description |
| --- | --- |
| sex | Sex of patient |
| age | Patient's age |
| leukocytes | Leukocytes counts (per $\mu\text{L}$) |
| neutrophilsP | Neutrophils percentage (%) |
| lymphocytesP | Lymphocytes percentage (%) |
| monocytesP | Monocytes percentage (%) |
| eosinophilsP | Eosinophils percentage (%) |
| basophilsP | Basophils percentage (%) |
| neutrophils | Neutrophils counts (per $\mu\text{L}$) |
| lymphocytes | Lymphocytes counts (per $\mu\text{L}$) |
| NLR | Neutrophil/Lymphocyte ratio |
| monocytes | Monocytes counts (per $\mu\text{L}$) |
| eosinophils | Eosinophils counts (per $\mu\text{L}$) |
| basophils | Basophils counts (per $\mu\text{L}$) |
| redbloodcells | red blood cells counts (per $\mu\text{L}$) |
| mcv | Mean corpuscular volume (fL) |
| mch | Mean corpuscular hemoglobin (pg) |
| mchc | Mean corpuscular hemoglobin concentration (mg/dl) |
| rdwP | Red Blood Cell Distribution Width (%) |
| hemoglobin | Hemoglobin (mg/dl) |
| hematocritP | Hematocrit percentage (%) |
| platelets | Platelets ($\text{mm}^3$) |
| mpv | Mean platelet volume |
| Condition | Positive or negative results for Covid-19 |

### 3.2 Demographic variables

Table 2 shows some summary statistics of the demographic variables (sex and age) before further processing:

**Table 2:** Demographic data summary for ECU raw dataset.

| Dataset | Samples | PCR Positive |  |  | PCR Negative |  |  |
| --- | --- | --- | --- | --- | --- | --- | --- |
| ECU | Selected | Male | Female | Total | Male | Female | Total |
| Sample size | 375 | 75 | 49 | 124 | 160 | 91 | 251 |
| Mean age | 36.86 | 38.41 | 35.00 | 37.06 | 37.09 | 36.18 | 36.76 |
| Standard deviation | 12.84 | 14.22 | 14.33 | 14.3 | 11.26 | 13.46 | 12.08 |

Over all, we can see that 62.67% of the clinical cases are males while 37.33% of the clinical cases are females. Also, 33.07% of these clinical cases test positive while 66.93% of these clinical cases test negative.

### 3.3 Blood variables

Some boxplots of all the blood variables before further processing are shown in Figure 2 in their original scale:

**Figure 2:**
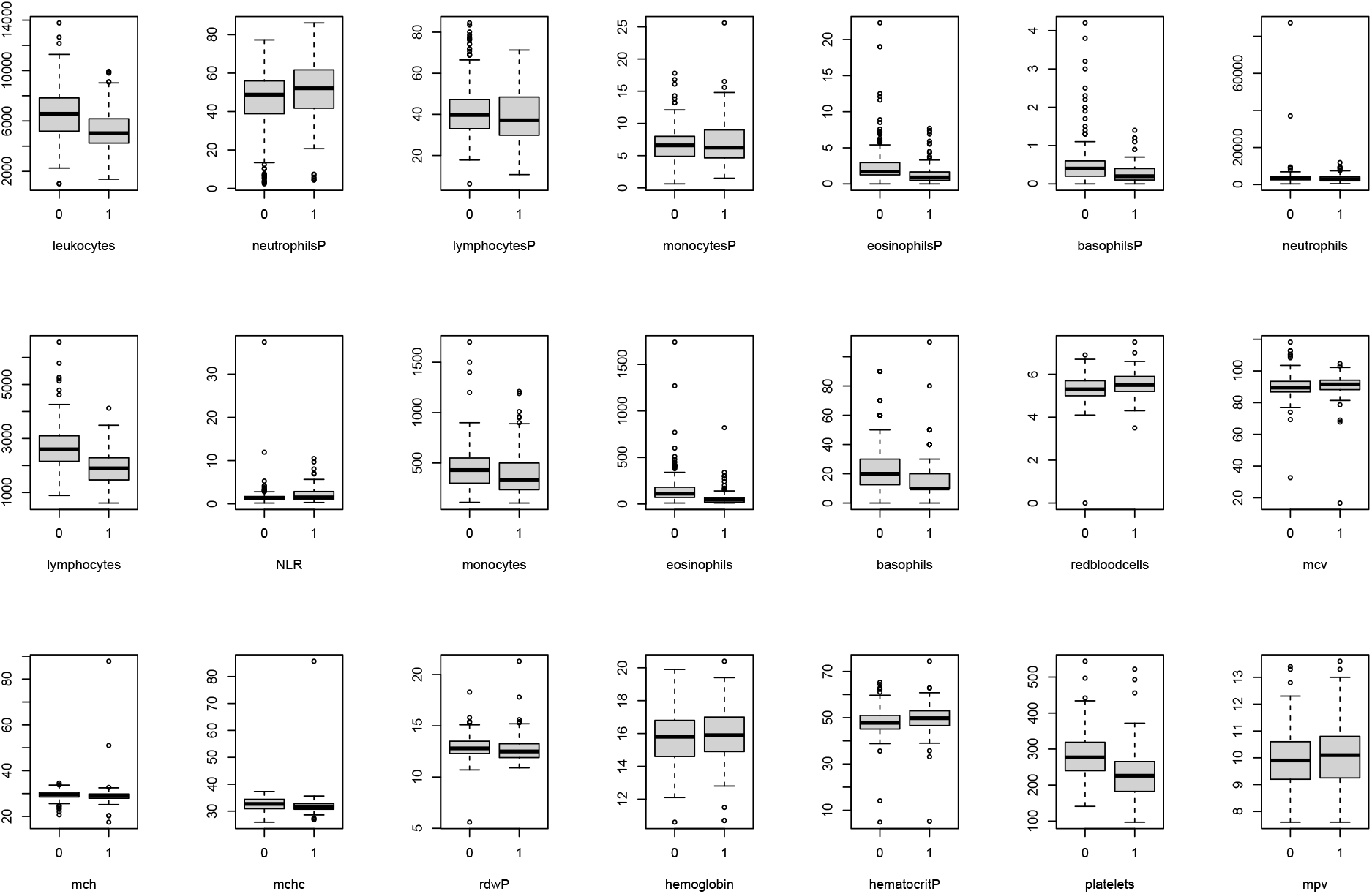
Boxplots of all the blood variables across different condition groups.

From Figure 2, we can see that some variables are positively-associated with the COVID condition, including neutrophilsP and mpv. These variables have relatively higher levels in the positive compared to the negative. Also, we can see that some variables are negatively-associated with COVID condition, including leukocytes, lymphocytes, eosinophils, basophils, platelets, etc. These variables have relatively higher levels in the negative group compared to the positive group.

### 3.4 Data cleaning

In the dataset, we find out that the leukocytes counts data do not match the summation of five WBC counts data. This problem cannot be fully explained by the presence of other white blood cells such as plasma cells, as we observe the summation of five WBC counts data exceeds leukocytes counts data in some cases (see the top of Figure 8 and Figure 9 in Supplementary Material). The presence of a large deviation of between the sum and the leukocyte counts indicates that special medical conditions may have happened. For example, certain viral infections can cause a decrease in circulating lymphocytes, while an intense allergic reaction can result in a significant increase in eosinophils. These changes can cause discrepancies in the total numbers. In the case of this data, all the patients were symptomatic and we want to exclude those cases for an accurate compositional data transformation.

Based on the reasons given above, we first apply some data cleaning procedure to our dataset. Patients whose leukocytes counts and WBC sum have a relative deviance larger than 10% were excluded, and the updated summary statistics are shown in Table 3. Updated deviance figures can be found in Supplementary Material (see the bottom of Figure 8 and Figure 9).

**Table 3:** Data Summary for ECU Cleaned Dataset.

| Dataset | Samples | PCR Positive |  |  | PCR Negative |  |  |
| --- | --- | --- | --- | --- | --- | --- | --- |
| ECU | Selected | Male | Female | Total | Male | Female | Total |
| Sample number | 329 | 65 | 42 | 107 | 146 | 76 | 222 |
| Mean age | 37.16 | 39.49 | 36.19 | 38.2 | 36.92 | 36.16 | 36.66 |
| Standard deviation | 12.79 | 13.96 | 14.46 | 14.18 | 11.29 | 13.49 | 12.07 |

It calls to the attention that 67.48% of the clinical cases is PCR-negative in our apparent dataset, whereas all of them were symptomatic before taking the blood test and PCR test. This issue of low-sensitivity of the PCR test will be discussed in greater details in the discussion and conclusions section.

Another thing to notice is that our dataset contains redundant information. For example, we are given the cell counts data of neutrophils, lymphocytes, monocytes, eosinophils, basophils and their summation data along with their percentages. Also, we are given the neutrophil/lymphocytes ratio data (NLR) despite the fact that we can calculate these values directly. Based on this observation, some of our variables should be excluded before we perform preliminary PCA on all possible predictors and try to avoid collinearity problems. The sex variable is a categorical variable that we should exclude in our PCA as well.

### 3.5 Ethics Statement

The review committee approved the study at the Technical University of Manabí with protocol number CBI-UTM-20-11-20-ROA. Every Laboratory informed the study motivation and obtained patients’ willingness before practicing the test. In the case of underage participants, informed consent was obtained from parents or legal guardians.

## 4 Data Analysis

In this section, we apply PCA, LDA, LR-PCA, and LR-LDA to the Ecuador CBC dataset and assess their performances.

### 4.1 PCA

#### 4.1.1 PCA using all variables

Before the principal component analysis, we exclude the following variables because of redundant information issue mentioned earlier: leukocytes, NLR, and percentage variables of five major WBC. Moreover, gender is not used for the PCA as it is binary in our dataset. The condition variable (RT-qPCR) is also excluded as it is our target outcome. Employing the PCA technique to all the other variables of the Ecuador CBC data gives us a biplot of Figure 3A. This biplot excludes one extreme point/outlier, in order to get a reasonable visualization of all the other observations.

**Figure 3:**
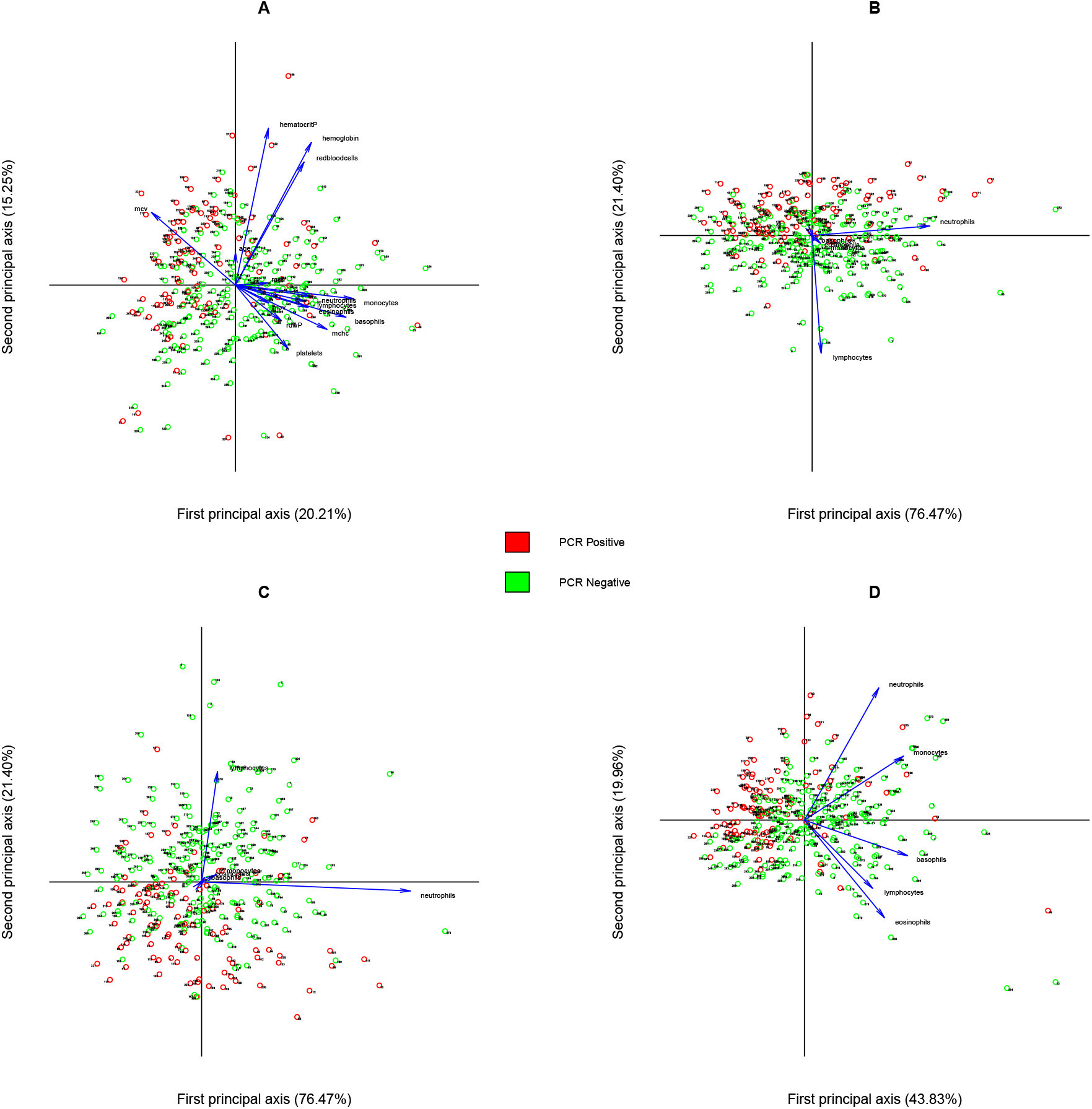
PCA biplots panel. A: Form biplot of PCA using all variables; B: Form biplot of PCA using WBC; C: Covariance biplot of PCA using WBC; D: Correlation-based Form biplot of PCA using WBC.

In the biplot, the cases with COVID-19 positive condition are colored as red points while the cases with COVID-19 negative condition are colored as green points. We use the correlation matrix in our PCA since we have many variables with incomparable units. Also, we use the form biplot for final demonstration. According to this data-visualization, our first principal component seems to be an important feature for COVID-19 condition classification, as most of the positive cases are located on the left hand side of our figure while negative cases are located on the right. This component is negatively associated with the mcv variable and positively associated with all the other variables. However, the first two principal components only explain 35.46% of the total variance, suggesting only using two principal components may be insufficient to capture most variance of our dataset. We need eight principal components for our selected features if we aim to explain more than 80% of total variance.

#### 4.1.2 PCA using WBC counts variables

Considering the prior knowledge of the diagnostic power of the five major WBC counts, we reemploy the PCA technique to the WBC counts dataset to give us a second form biplot (see Figure 3B). In this analysis, we only include neutrophils, lymphocytes, monocytes, eosinophils and basophils cell counts as our predictor variables.

We are using the covariance-based PCA first since we are dealing with count variables and their units are perfectly comparable with each other. According to this data-visualization, the second principal component seems to be an important feature for COVID-19 condition classification, as most of the positive cases are located at the top of the figure while negative cases are located at the bottom. And this component is highly correlated with the lymphocytes variable and almost uncorrelated with all the other variables. Moreover, the first two principal components already explain 97.87% of total variance, suggesting using two principal components is sufficient to capture most of the white cell count dataset’s structure. One noteworthy observation is that there still exists mixing of positive and negative cases (e.g. cases 40,77,160, and 188) since the PCA method is not optimized for class separation. This mixing may be attributed to factors such as the presence of other respiratory diseases or variations in the stages of the immunological window. Further details are discussed in the Discussion and Conclusions section. Moreover, we can plot a covariance biplot (also based on the covariance matrix instead of correlation matrix) to visualize the correlation structure of the variables (see Figure 3C). From both the form biplot and the covariance biplot, we can see that lymphocytes and neutrophils dominate the first two PCs. They have the largest counts and also the largest variances. To reduce the influence of the large count number of lymphocytes and neutrophils, we could also standardize the data and make a correlation based form biplot as shown in Figure 3D. In this correlation based form biplot, the first two principal components only explain 63.80% of the total variance.

From the correlation-based biplot, we can see that now five counts variables have similar length and all five of them are positive correlated with the first principal component. Since most of PCR positive cases are located on the left half of the biplot and most PCR negative cases are located on the right half of the biplot, it seems that all five WBC counts variables are positively correlated with the PCR negative status in our standardized WBC counts dataset. Thus, in this analysis, we can interpret the first principal component as the total WBC count and the second principal component as the neutrophils-eosinophils difference, with the total count having a high predictive power for COVID-19 status classification. This information is not evident from the covariance-based biplots, as lymphocytes and neutrophils dominate the first two PCs.

### 4.2 LR-PCA

Now we apply LR-PCA to the WBC counts data. In order to do so, the WBC counts data is first transformed to a compositional dataset by dividing each entry with its row summation (the sum of that cases’ WBC counts). Then we are able to apply the *clr* transformation followed by a standard PCA to obtain a form biplot (see Figure 4A). Notice that we have eight cases with zero basophils counts in our WBC counts dataset, and these zeros are replaced with a small non-zero number based on a Bayesian-multiplicative replacement by the cmultRepl function in the zCompositions R package [14].

**Figure 4:**
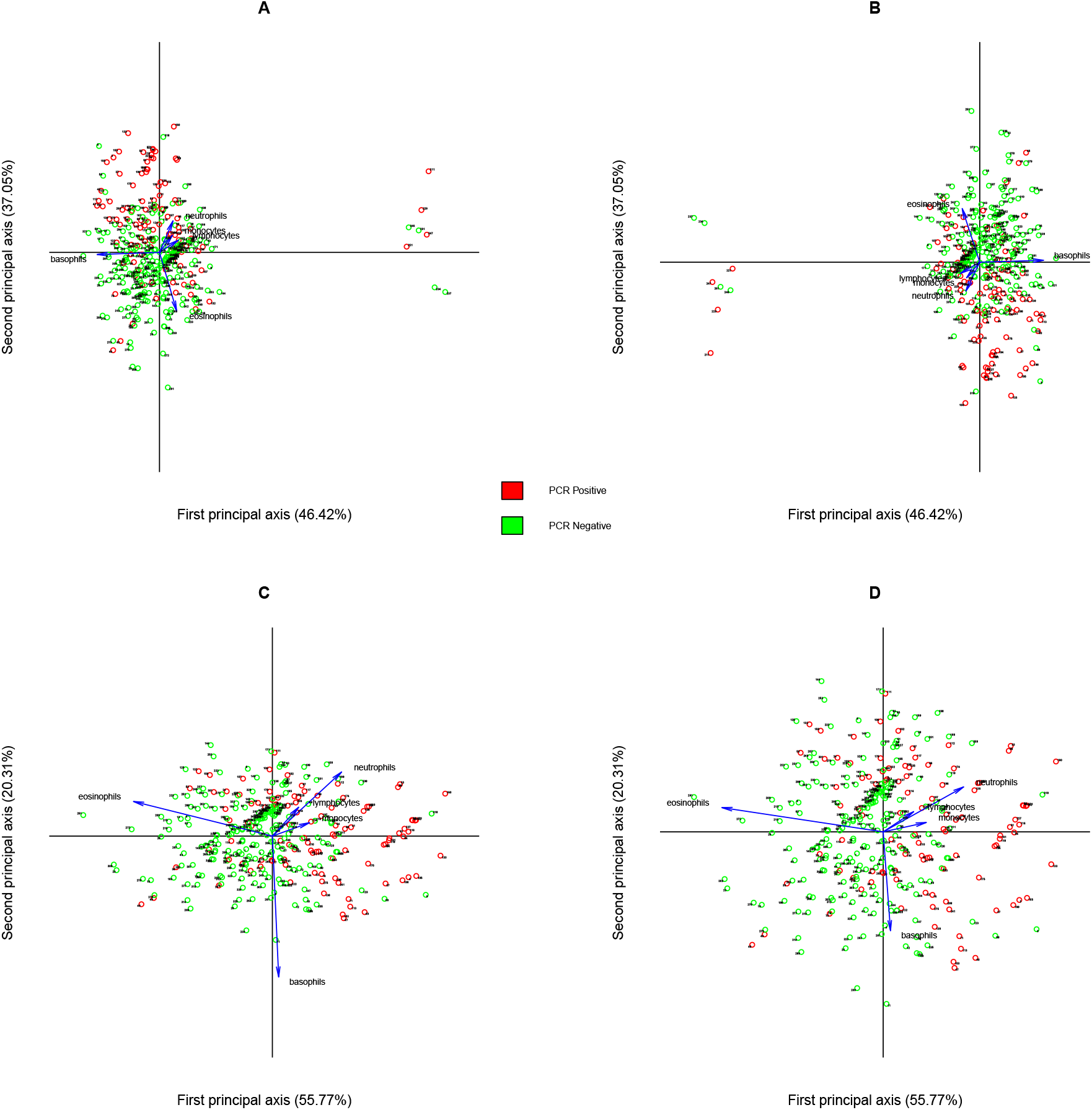
LR-PCA biplots panel. A: Form biplot of LR-PCA using WBC; B: Covariance biplot of LR-PCA using WBC; C: Form biplot of LR-PCA using WBC removing outliers; D: Covariance biplot of LR-PCA using WBC removing outliers.

According to this data-visualization, the log-ratio of basophils and lymphocytes almost coincides with the first principal axis, and the log-ratio of eosinophils and neutrophils almost coincides with the second principal axis. Moreover, the log-ratio of eosinophils and neutrophils is almost uncorrelated with the log-ratio of basophils and lymphocytes, as the links between those vectors has an angle close to 90 degrees. In the form biplot, the first two principal components explain 83.47% of the total variance, suggesting using two principal components is sufficient to capture most of *clr* -transformed WBC composition dataset’s structure. One thing to notice is that the minimum eigenvalue of the sample covariance matrix is systematically zero, and this is caused by the unit-sum constraint of the compositional data. Also, we have eight extreme points in the biplot, and these coincide with eight cases having zero basophils counts. Moreover, we can plot a covariance biplot (also based on covariance matrix) to visualize the correlation structure of *clr* -transformed WBC composition dataset (see Figure 4B). Still, we have eight extreme points in the biplot, corresponding to eight cases with zero basophils counts. From both the form biplot and covariance biplot, we can see that the log-ratio of basophils and lymphocytes almost coincides with the first principal axis, and the log-ratio of eosinophils and neutrophils almost coincides with the second principal axis. Moreover, the log-ratio of eosinophils and neutrophils is almost uncorrelated with the log-ratio of basophils and lymphocytes. These aspects are overlooked if we only visualize our dataset by naive PCA and only captured if we view our dataset as a compositional dataset.

Figure 3C and Figure 3D are the form biplots and covariance biplots of the WBC compositional dataset if we remove the eight basophils count zero cases. Notice that the direction of basophils and eosinophils vectors changes a lot if we remove the effect of those outliers. Once the outliers are removed, the eosinophils/neutrophils appears as the most variable ratio in the dataset. However, as we can see in the next section, removal of the outliers hardly affects the classification results as the change hardly affects the LR-LDA coefficients. Therefore, we only include those biplots for reference purposes and still use the processed WBC compositional dataset (329 cases) that includes the eight outliers.

### 4.3 LDA

#### 4.3.1 LDA using all variables

We apply LDA to the full dataset and WBC counts dataset before the log-ratio transformation. Figure 5A shows the group separation obtained with the *standardized* full dataset. Notice that standardization won’t give us different classification results because of the invariance property of LDA, and we standardize our full dataset in order to get comparable linear discriminant coefficients for all variables.

**Figure 5:**
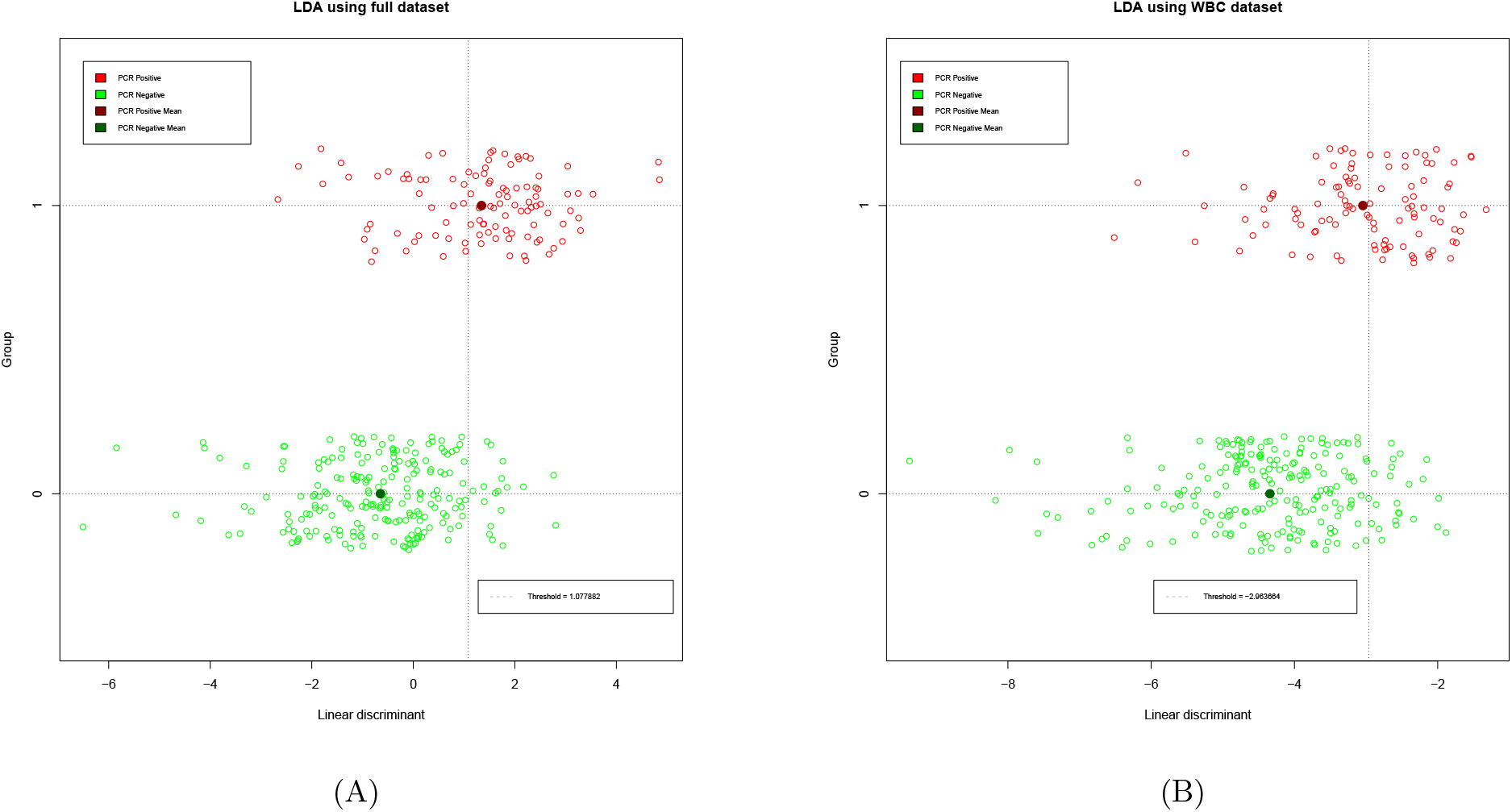
Combined LDA visualizations. A: LDA using all variables; B: LDA using five major WBC counts variables.

We add a small random noise (through the “jitter” function in R) to the COVID condition variable prior plotting the results to reduce the amount of overlapping points. In Figure 5A, we are classifying the outcome without cross-validation to obtain a classifier threshold 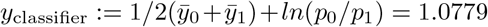 (plotted as the vertical dotted line, with equal cost for false positives and false negatives). Data points on the left hand side of the classifier are classified as PCR negative and data points on the right hand side are classified as PCR positive. From Figure 5A we can see that the LDA technique performs better for classifying PCR negative patients compared to PCR positive patients, as more red points are misclassified to the left hand side of the classifier compared to green points on the right hand side of the classifier. This fact can also be seen by the confusion matrix of LDA on the full dataset (see Table 6, with 18 *<* 36). We obtain a classification rate of 83.587% using LDA without cross-validation.

We present the coefficient vector **a** of linear discriminant function *y*_*i*_ = **a**^*T*^ · **x**_*i*_ in Table 4. In Table 4, the variables with a positive coefficient are variables positively associated with the COVID-19 condition, while those variables with a negative coefficient are negatively associated with the COVID-19 condition. From both the table and the bar plot, we can see that the variable lymphocytes has the most prediction power. Red blood cells, platelets, and eosinophils also have relatively large prediction power, as they have coefficients with a large absolute value. According to the linear discriminant function coefficient, mcv doesn’t have much more prediction power compared to other variables and this observation is different from the the PCA biplot using all variables shown in Figure 3A. The reason behind this inconsistency could be due to the different targets of PCA and LDA techniques. Surprisingly, the monocytes count variable has a different direction compared to all the other four WBC count variables (neutrophils, lymphocytes, eosinophils and basophils) in the LDA. However, in the principal component analysis all five WBC counts variables have similar directions (see Figure 3B). This inconsistency could be due to several reasons that need further investigation. For example, the reason could be statistic. The compositional data structure requires WBC counts variables to sum up to their total counts, and this collinearity problem could result in the constraint of their LDA coefficient making the direction of monocytes variable to flip. Also, some hidden biological mechanisms linking monocytes counts variable with the COVID-19 condition may cause the sign of monocytes to flip when it comes to COVID-19 prediction instead of just data visualization.

**Table 4:**
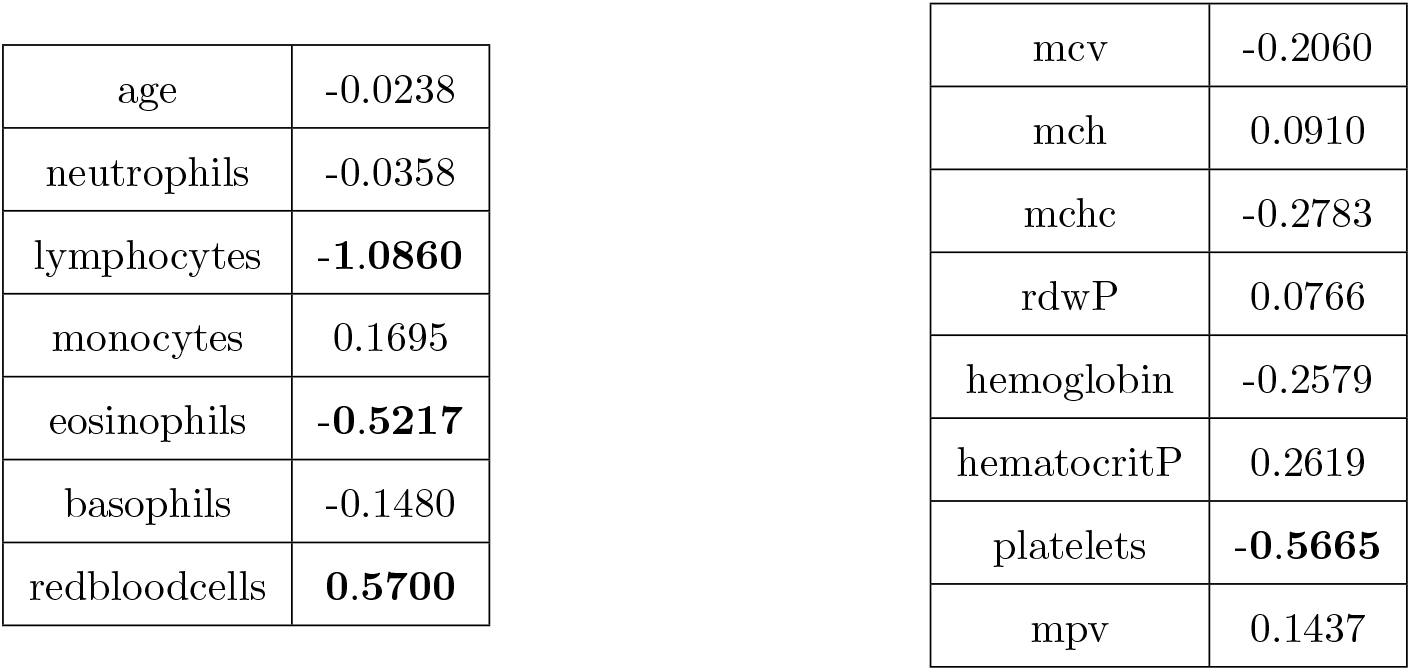
Coefficient vector of the linear discriminant function (normalized full dataset).

The receiver operating characteristic curve (ROC curve) for LDA using all variables is shown in the combined ROC Figure 7 (labeled as LDA Full). We use the roc function from the R package pROC to generate all the ROC figures in this paper[20]. The area under the curve (AUC) is 0.8487 without cross-validation. The classification rate is calculated using leave one out (LOO) cross validation. We obtain a classification rate of 81.459% with the corresponding confusion matrix (see Table 6)

#### 4.3.2 LDA using WBC counts variables

We repeat the LDA analysis using only the WBC counts. Figure 5B demonstrates LDA with the WBC dataset. Since all five WBC counts variables have the same unit, there is no need to standardize our dataset this time.

We continue to use a small random noise to the COVID condition variable prior plotting the results to reduce the amount of overlapping points. In Figure 5B, we are classifying outcome without cross-validation to obtain a classifier threshold 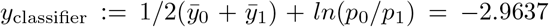 (plotted as the vertical dotted line, with equal cost for false positives and false negatives). Data points on the left hand side of the classifier are classified as PCR negative and data points on the right hand side are classified as PCR positive. From Figure 5B, we can see the LDA technique still performs better for classifying PCR negative patients compared to PCR positive patients in WBC dataset, as more red points are misclassified to the left hand side of classifier compared to green points on the right hand side of classifier. Moreover, the LDA technique performs worse in the WBC dataset compared to the full dataset as we have more misclassified cases. Those facts can also be seen by the confusion matrix of LDA on the WBC dataset (see Table 6). We obtain a classification rate of 77.508% using LDA without cross-validation.

We present the coefficient vector **a** of linear discriminant function *y*_*i*_ = **a**^*T*^ · **x**_*i*_ for the WBC dataset in Table 5. Again, we have variables with a positive coefficient sign indicating that those variables are positively associated with the COVID-19 condition, while variables with a negative coefficient sign indicate that they are negatively associated with the COVID-19 condition. From both the table and the bar plot, we can see that the basophils variable has the most prediction power. This observation is hard to see in the PCA biplot Figure 3B as well, and could be due to the different targets of PCA and LDA techniques.

**Table 5:** Coefficient vector of the linear discriminant function (WBC dataset).

|  |  |
| --- | --- |
| neutrophils | -0.0001 |
| lymphocytes | -0.0014 |
| monocytes | 0.0010 |
| eosinophils | -0.0025 |
| basophils | **-0.0137** |

**Table 6:** Combined performance summary for LDA models.

| Models | TN | FP | FN | TP | Classification Rate | AUC |
| --- | --- | --- | --- | --- | --- | --- |
| LDA full without CV | 204 | 18 | 36 | 71 | 83.587% | 0.8487 |
| LDA full with CV | 200 | 22 | 39 | 68 | 81.459% | 0.8159 |
| LDA WBC without CV | 199 | 23 | 51 | 56 | 77.508% | 0.8129 |
| LDA WBC with CV | 198 | 24 | 52 | 55 | 76.900% | 0.7989 |

The receiver operating characteristic curve (ROC curve) for LDA using WBC counts variables is shown in the combined ROC Figure 7 (labeled as LDA WBC). The area under the curve (AUC) is 0.8129 without cross-validation. The classification rate is also calculated using LOO cross validation. We obtain a classification rate of 76.900% with the corresponding confusion matrix (see Table 6). Table 6 summarizes the performances of LDA classification models using all variables and WBC counts variables with or without cross-validation. The classification rates are more accurate with cross-validated models as they compensate for overfitting problems.

### 4.4 LR-LDA

We apply the LR-LDA method by applying the function lrlda in the R package ToolsForCoDa [6] to our compositional dataset, and we obtain Figure 6.

**Figure 6:**
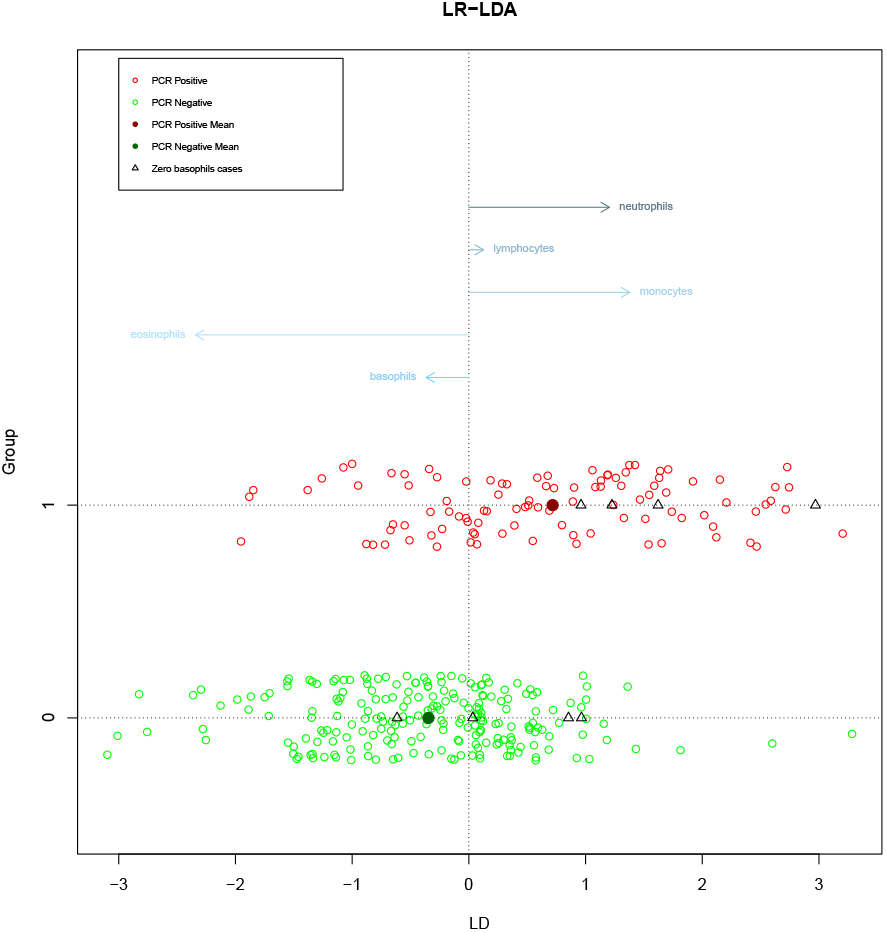
LR-LDA of WBC compositional data showing classification results and coefficient vectors.

**Figure 7:**
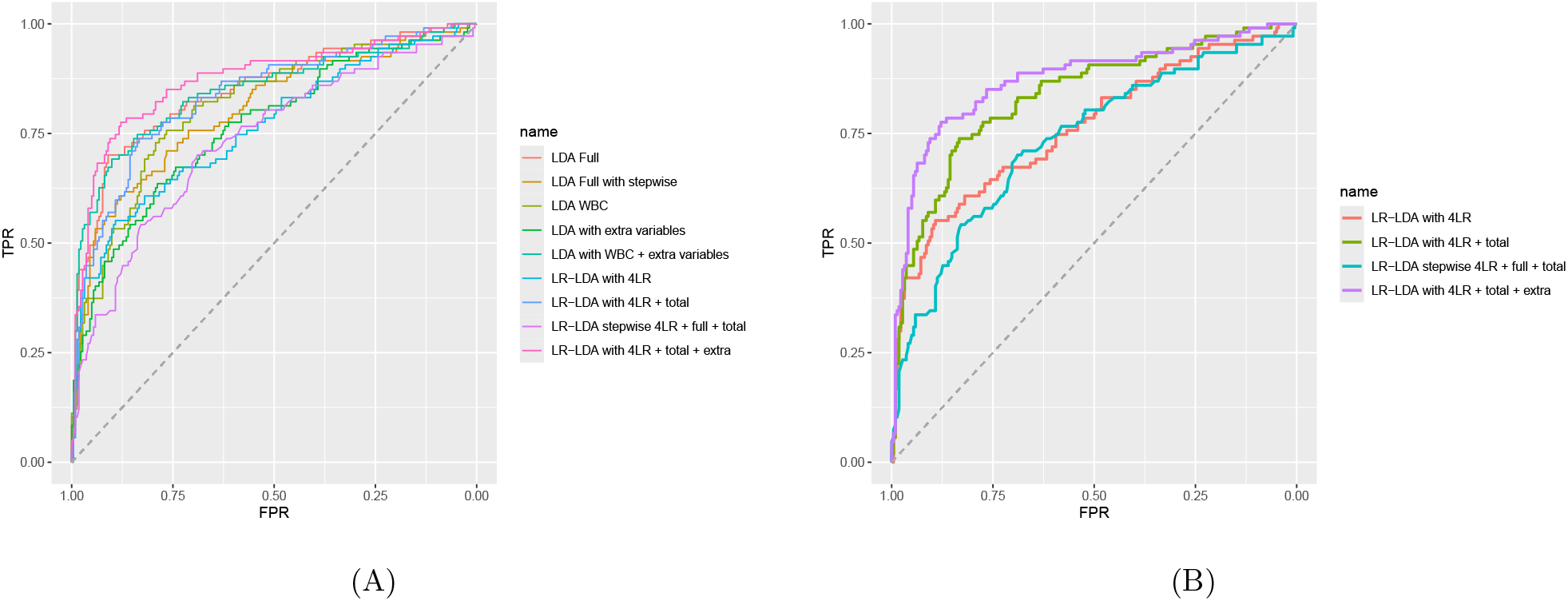
The ROC curves. A: The ROC curves for all models; B: The ROC curves for compositional models.

**Figure 8:**
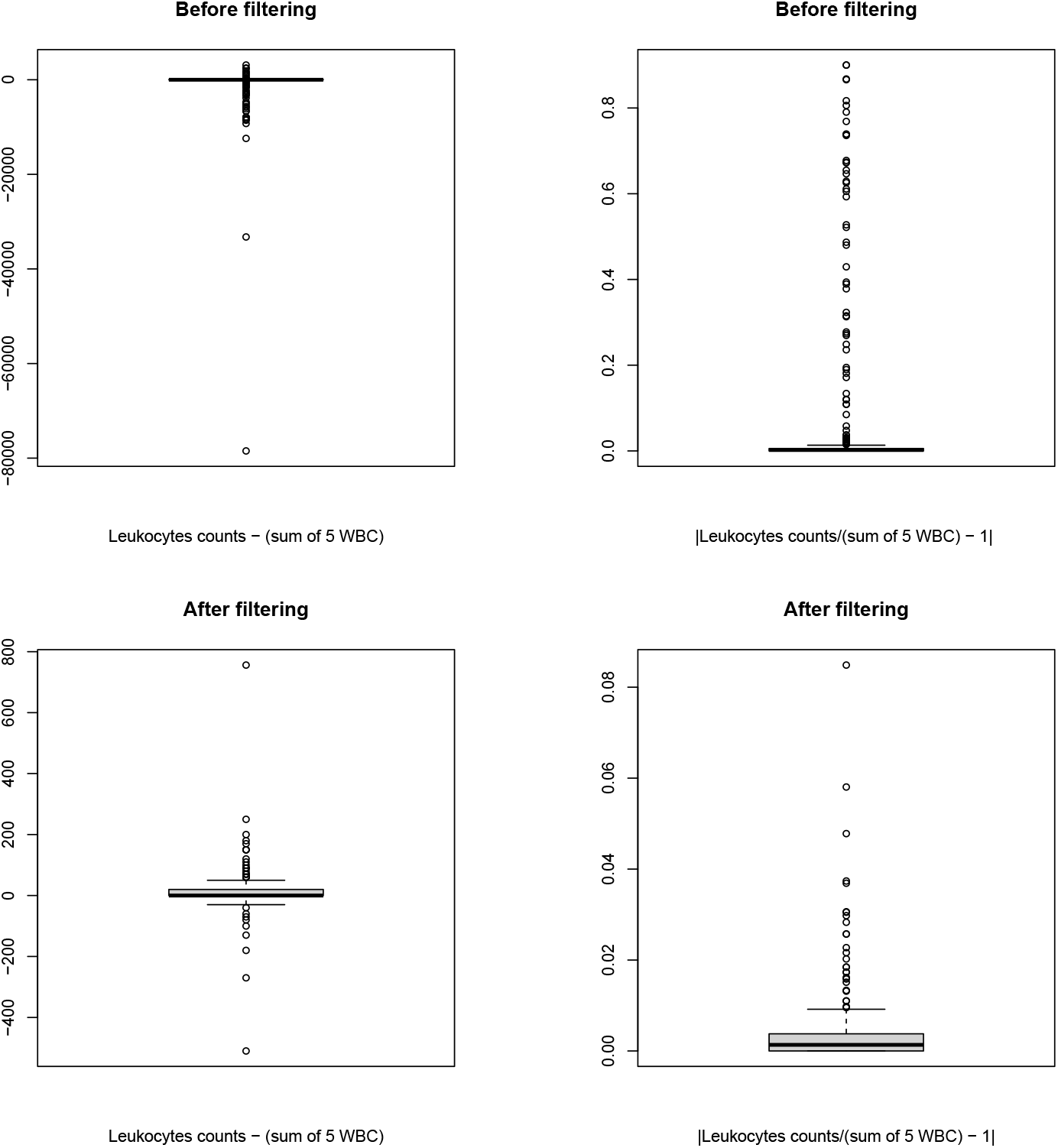
Boxplots of absolute and relative deviance before and after filtering.

**Figure 9:**
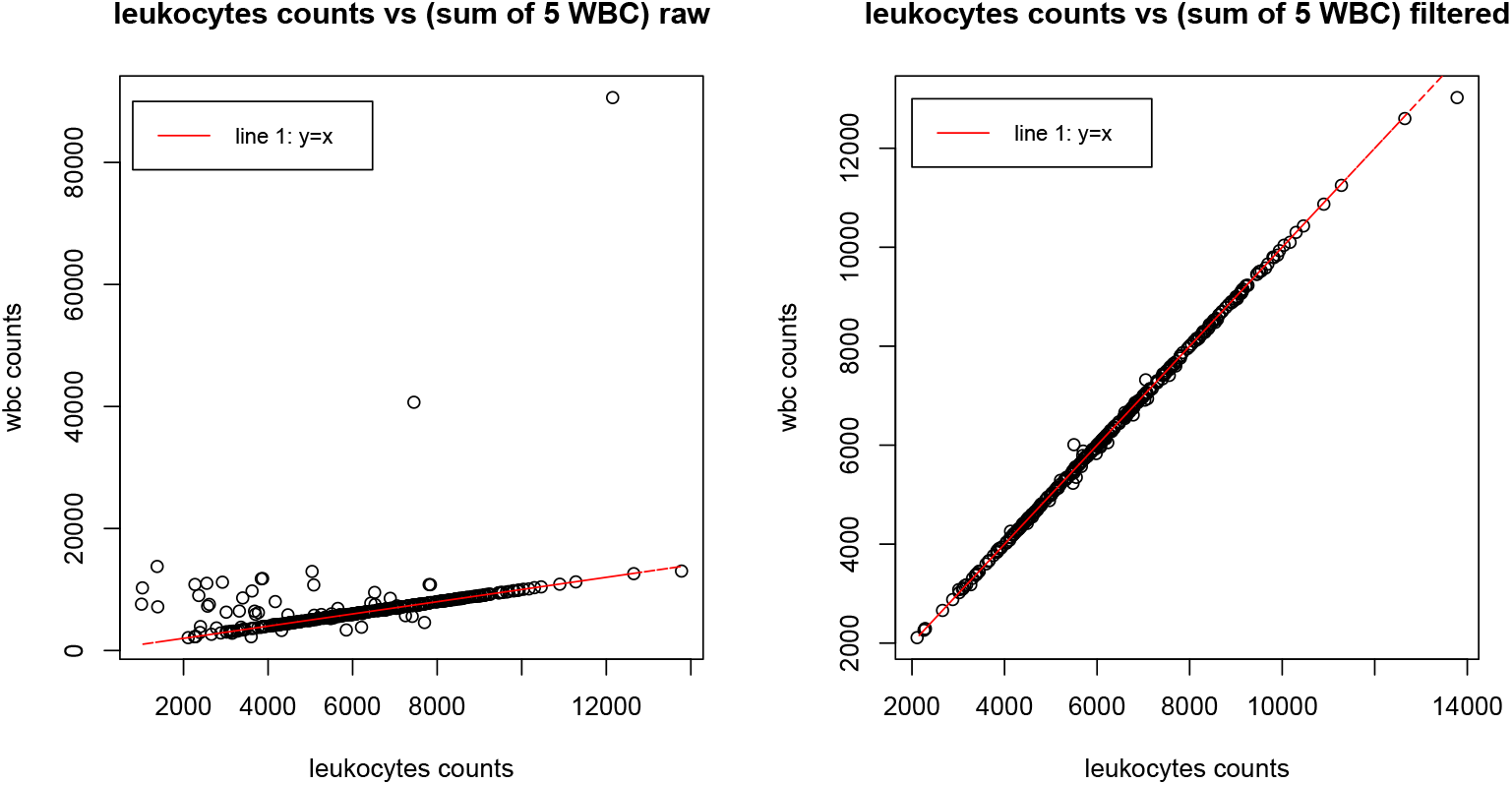
Scatterplots of WBC summation vs leukocytes counts before and after filtering.

We again add a small random noise to the COVID condition variable to avoid overlapping. The arrows on the top of Figure 6 represent the coefficients obtained in the LR-LDA, it is one dimensional since we are only dealing with two groups. Also, we represent the eight cases with zero basophils as triangles and noticed that they are no longer outliers in the LR-LDA analysis. Similar to the linear coefficient of the LDA analysis part, larger distance between vector arrows can be interpreted as more powerful log-ratios when predicting the COVID-19 condition outcome. Notice that the LR-LDA technique performs a little worse in the WBC composition data compared to the WBC counts dataset as we have more misclassified cases. This fact can be seen in the confusion matrix of LR-LDA in Table 7A. Moreover, we obtain a classification rate of 77.51% using LR-LDA. A visualization of LR-LDA vectors (coefficients) can be seen in Figure 6 and Table 7A.

**Table 7:**
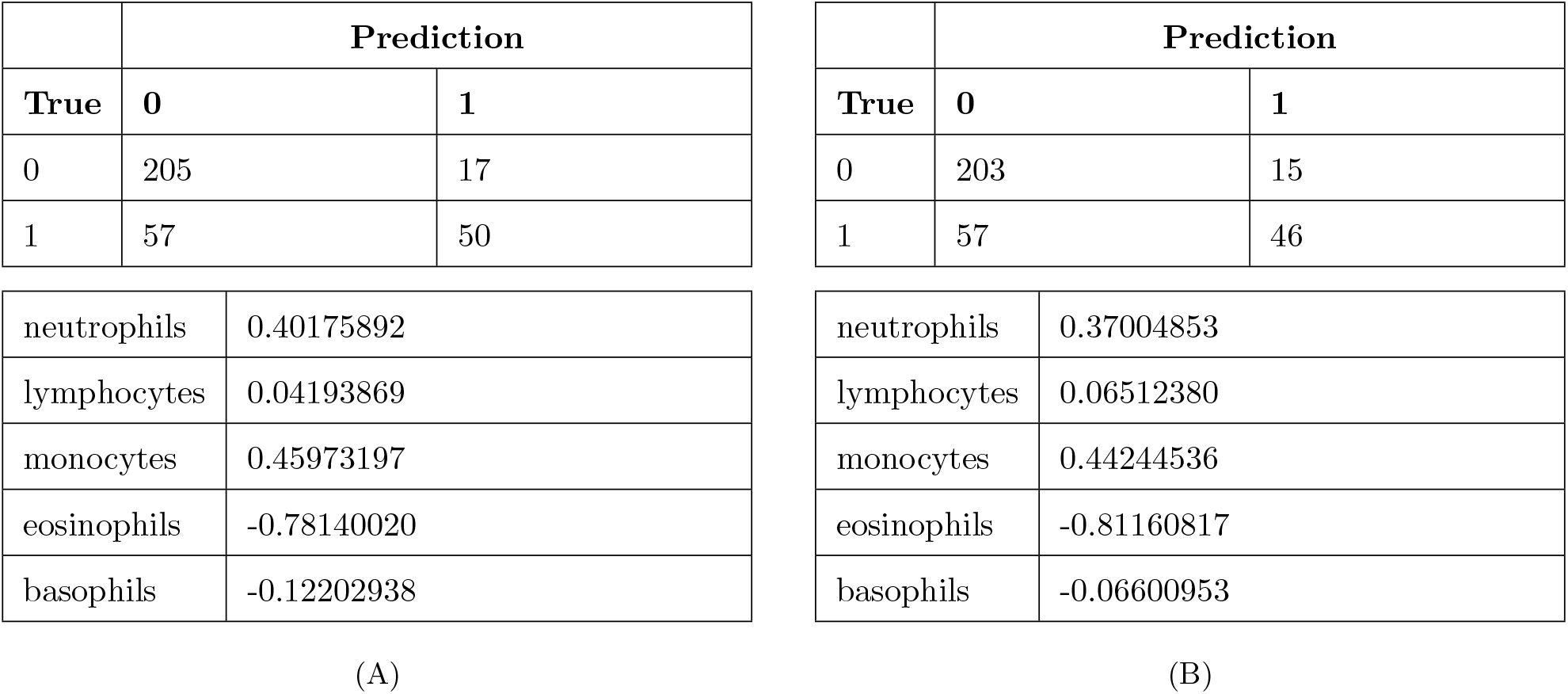
Combined results for LR-LDA with WBC composition data. A: LR-LDA with WBC composition data; B: LR-LDA with WBC data and eight outliers removed.

From the coefficients vectors in Figure 6 and Table 7A, we can see that the log-ratio of neutrophils and eosinophils is one of the most discriminating log-ratios between the variables (only slightly less than the log-ratio of monocytes and eosinophils), as they have a relative large distance between the arrows (0.40 − (−0.78) = 1.18). This observation also coincides with our LR-PCA biplots (both the form biplot and the covariance biplot), as the log-ratio of neutrophils and eosinophils almost coincides with the second principal axis which has a relative large classification power of PCR positive cases and PCR negative cases. Moreover, the contrast of neutrophils and eosinophils also appeared in the bottom right of Figure 3, where their difference almost coincides with the second principal axis and explains most of the data structure together with the total WBC counts (which coincides with the first principal axis). The ROC curve for the LR-LDA method is shown in the ROC Figure 7 (labeled as LDA with 4LR). The area under the curve (AUC) is 0.7625. If we remove those eight outliers instead, we have the following confusion matrix and coefficient vector of the LR-LDA. Now the classification accuracy is 77.57% instead of 77.51%. We can see the difference of the classification results and coefficient vectors by comparing Table 7A with Table 7B. As we mentioned at the end of LR-PCA section, removing eight outliers hardly affects the classification results and the change doesn’t affect much of LR-LDA vector’s coefficient. Therefore, we keep the eight outliers in the analysis.

### 4.5 Feature selection

Applying LR-PCA and LR-LDA to the WBC compositional data did give us some extra insight of its structure, but they don’t give us a satisfactory classification rate that improves over the rate obtained with the full dataset LDA using all information. Therefore, we try to add some extra variables to the compositional dataset hoping to achieve a better classification rate. Notice that just adding the clr transformed cell counts to the full dataset will cause a singularity problem as the clr transformed counts are systematically collinear. Therefore, we apply the ilr-transformation as in Equation (10) (instead of clr as in Equation (8)) to our WBC counts dataset to avoid the collinearity problem in the whole feature selection section. Now our WBC compositional dataset is represented by four log-ratios. We note that without additional non-compositional predictors, the classification results and AUC scores will be exactly the same using ilr-transformation and clr-transformation, since both methods give exactly the same posterior probabilities.

#### 4.5.1 Adding total WBC counts

In compositional data analysis, the total number of all the components is often ignored. Although in CoDA we only care about the relative values in our dataset, their total number, which is the total number of white blood cell counts can be a powerful predictor for the outcome COVID-19 infection status. Therefore, we calculate the summation of WBC counts of each patient and treat it as a new variable. Then we include this total WBC counts variable into our WBC compositional dataset and apply LR-LDA method. Since now we have a non-singular covariance matrix as we choose to use ilr-transformation for feature selection, specific programs that use generalized inverses are no longer needed and the analysis can be performed with the standard R function lda.

The results for the LR-LDA classification using ilr-transformed WBC composition data plus the total WBC counts variable are presented in the Table 8. The classification accuracy is 79.64% before cross-validation. This result is slightly better than the LR-LDA classification itself but still not as good as the LDA classification using the full dataset. The ROC curve of using ilr-transformed WBC composition data plus the total WBC counts variable is presented in the Figure 7. This analysis shows that complementing the log-ratio transformed compositions with their total count can be helpful.

**Table 8:** Combined summary of all models with confusion-matrix entries (TN, FP, FN, TP) after CV.

| Model | Acc (bef. CV) | AUC | Acc (aft. CV) | TN | FP | FN | TP |
| --- | --- | --- | --- | --- | --- | --- | --- |
| LDA Full | 83.59% | 0.8487 | 81.46% | 200 | 22 | 39 | 68 |
| LDA Full with step-wise | 80.55% | 0.8028 | 80.55% | 209 | 13 | 51 | 56 |
| LDA WBC | 77.51% | 0.8129 | 76.90% | 198 | 24 | 52 | 55 |
| LDA with extra variables | 76.60% | 0.7649 | 75.08% | 201 | 21 | 61 | 46 |
| LDA with WBC + extra variables | 82.98% | 0.8456 | 82.07% | 200 | 22 | 37 | 70 |
| LR-LDA with 4LR | 77.51% | 0.7625 | 76.90% | 204 | 18 | 58 | 49 |
| LR-LDA with 4LR + total | 79.64% | 0.8354 | 79.03% | 200 | 22 | 47 | 60 |
| LR-LDA stepwise 4LR + full + total | 73.56% | 0.7384 | 73.56% | 209 | 13 | 74 | 33 |
| **LR-LDA 4LR + total + extra** | **85.11%** | **0.8727** | **84.50%** | 206 | 16 | 35 | 72 |

#### 4.5.2 Step-wise LDA

Another common approach for feature selection is to use a step-wise algorithm. We use the train function with “method = stepLDA” from the R package caret to apply step-wise LDA automatically [9]. In the train function, a grid of all parameters is created and the model is trained on slightly different data for each candidate combination of tuning parameters. Across each data set, the performance of held-out samples is calculated and the mean and standard deviation is summarized for each combination. The combination with the optimal resampling statistic (in our case this is the classification accuracy) is chosen as the final model and the entire training set is used to fit a final model[9]. When we fit the Step-wise LDA model, we use the full dataset excluding the five WBC counts variables, and include the ilr transformed compositional variables (four of them) and the total WBC counts variable. With a 10-fold cross validation while selecting variables, the train function gives us the final model as:

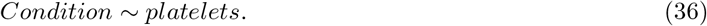

The classification accuracy for this model is 73.56% before cross-validation. This result is actually worse compared to the LR-LDA classification and the LDA with WBC composition data adding total WBC counts. The confusion matrix is presented in the Table 8 below. The ROC curve of using step-wise LDA model is presented in the Figure 7.

Also, we can fit the Step-wise LDA model over the full dataset without adding compositional variables and the total WBC counts variable, to make our result comparable to the full dataset LDA. The train function give us the final model as:

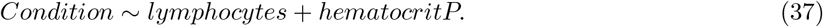

The classification accuracy for this model is 80.51% before cross-validation, which is worse compared to the naive LDA with the full dataset but gives a more parsimonious model. Its confusion matrix is presented in Table 8 below and its ROC curve is presented in the Figure 7.

#### 4.5.3 Adding predictive variables

Even the classification accuracy of the first model in this section (compositional dataset plus total WBC counts model) improved the naive LR-LDA compositional data model, it still fails to improve over the full LDA model which has a classification rate of 83.587% before cross-validation. However, the full LDA model has given us some insight of each variables’ prediction power, and we try to exploit this information by adding the most predictive variables other than WBC counts (i.e. variables with a large linear discriminant coefficient in absolute value in Table 4) and the total WBC counts variable to the compositional dataset and apply the LR-LDA method. The reason we do not add raw WBC counts variables is that their information should be fully incorporated in the compositional dataset as we view WBC counts as compositional variables and only care about their relative values.

As a result, when we add the red blood cells, the platelets and the total WBC counts variable to the ilr-transformed dataset and apply the LDA method to those seven variables, we obtain a classification accuracy rate of 85.11% before cross validation and a classification accuracy rate of 84.50% after cross validation. Both results are actually better compared to the best result we had before by using LDA on the full dataset (which has a classification rate of 83.58% before cross-validation and 81.46% after cross-validation). The after cross-validation confusion matrix is presented below in Table 8. The ROC curve is presented in Figure 7A. To highlight the ROC curves for compositional LDA models, a separate Figure 7B only including these ROC curves is also presented.

One thing to notice is that without the four compositional variables (given by ilr-transformation), simply applying the LDA method to the three added variables (red blood cells, platelets, and total WBC counts) will actually give us much worse results: classification accuracy of 76.60% before cross-validation and 75.08% after cross-validation. Also, if we just use the non-compositional LDA model with WBC counts variables adding predictive variables (red blood cells and platelets, no total WBC counts variables to avoid collinearity problem), we will also obtain worse results: classification accuracy of 82.98% before cross-validation and 82.07% after cross-validation. All of the results presented above are worse compared to the current compositional model, which means adding compositional variables to our LDA model does help us get a better understanding of data structure and helps us to improve the classification rate.

Table 8 that summarize our models and their performances so far are presented below.

## 5 Discussion and Conclusions

In our dataset, about 67% of the clinical cases is PCR-negative in our cleaned dataset, whereas all of them were symptomatic before taking the blood test and PCR test. The situation in which people with symptoms show negative PCR results is complex and can have several explanations.

We can think about a few scenarios that may have contributed to these results. The first reason for false negatives in PCR is related to the infection stage of patients and the different variants of the virus. The PCR test is most sensitive during the period of highest viral load, which usually occurs in the first few days after the onset of symptoms. If the tests were carried out later in the infection, the viral load may have decreased enough to generate a false negative. On the contrary, some patients who have been tested positive may be at the end of the immunological window while others at the beginning. In other words, the binary COVID-19 infection status variable is not enough to capture the continuous infection stage information, which make our classification attempt more complex and difficult. Also, the constant emergence of new variants of the virus during the pandemic, with their mutations, may have affected the sensitivity of the PCR test, leading to an increase in false negatives.

The presence of other pathogens, such as other respiratory viruses, and allergies can also lead to negative cases in the PCR, and those cases are not necessarily *false* negatives. For example, the symptoms may result from other common respiratory viruses, such as influenza or rhinovirus, circulating simultaneously with SARS-CoV-2. This is the most likely reason for most negative PCR results in the Ecuador database. Also, in some cases, symptoms such as runny nose, cough, and sore throat can be triggered by seasonal or other allergies, and not necessarily by a viral infection. Consequently, part of the presumably negative cases are, despite being symptomatic, in fact truly PCR-negative for they do not suffer from COVID-19, but from a different respiratory health problem.

There are also other factors that can contribute to this phenomenon. For example, The perception of symptoms can vary from person to person, and some individuals may report symptoms even in the absence of an infection. Also, the immune system can control the infection quickly in some cases, resulting in a low viral load and a negative PCR test, even in the presence of symptoms. Unfortunately, there is no single right answer. There are many possibilities and we need further investigation if we try to find out reasons behind both true and false negative cases.

The data analysis shows that PCA and LR-PCA can be useful for exploring the data. In particular, biplots are shown to be powerful for visualization for compositional, non-compositional, and mixed data. LDA and LR-LDA are shown to be efficient classification tools for categorical outcomes, and we are able to quantify the predictive power of each variable which turns out to be helpful for feature selection and potential interpretation. All computations and figure generations were completed using the statistical environment R [18], and the computational load was manageable on a laptop computer with 12th Gen Intel(R) Core(TM) i7-12700H 2.70 GHz processor and 16 Gigabytes RAM.

Moreover, we have seen that compositional data analysis methods (LR-PCA and LR-LDA) have given us some extra insights about the data structure and the best classification model for COVID-19 status prediction. To be more specific, from the LR-PCA visualization we have known that the log-ratio of neutrophils and eosinophils is a key feature for infection status classification. This information can not be obtained by the PCA with the full dataset or the PCA with the WBC counts dataset. And from the LR-LDA coefficients, we noticed that the log-ratio of neutrophils and eosinophils has relatively large classification power which confirms our observations again. By comparing the classification accuracy and the AUC of all models, we have shown that the compositional model with some extra variables has the best performance. This performance is due to the extra information we gained by adding compositional variables, as the model with only added variables or the model with the non-compositional WBC counts with extra variables performed worse compared to our compositional model.

To summarize, we have successfully illustrated the efficacy of combining compositional data analysis with traditional non-compositional data-visualization method and classification models. When our dataset has an intrinsic and scientifically meaningful compositional structure (like the WBC counts variables), we recommend combining compositional data analysis method with the classic analysis methods (like LR-LDA and LR-PCA) to gain extra insights and improve the performance of models. Moreover, we have successfully come up with a compositional classification model using only complete blood-count data. This compositional model satisfied our expectation to be a cheap and efficient tool to help diagnosing COVID-19 infection with a superior cross-validated accuracy. Notably, both compositional and non-compositional models consistently achieved higher classification accuracy for PCR negatives than for PCR positives in our dataset. From a practical point, in a scenario of limited availability of PCR tests, this result may be used to argue for a two step procedure, where the WBC test is used primarily to discard COVID-19 and the PCR test is applied only to those individuals that result positive in the WBC test. This may save considerable resources though some COVID-19 cases will go undetected.

Although in this paper we have been focusing on LDA and LR-LDA for classification models, other common methods (like logistic regression, K-nearest neighbor algorithm, Quadratic discriminant analysis, etc) could also be applied with compositional data and evaluated. Also, there are more advanced classification models like neural networks, support vector machines, LIME [19] and SHAP [12], and the inclusion of compositional information in the form of log-ratio transformed compositions using these methodologies is another interesting topic for future investigation.

The discussion above clearly suggests that the PCR-negative cases form a heterogeneous group, and this suggests a potential direction for future research. The situation of symptomatic patients showing negative PCR results is complex and has multiple explanations that are quite different from each other. We suggest the use of cluster analysis to potentially discover groupings in the negative cases. Inclusion of such a grouping may make our compositional classification model more accurate and interpretable.

Another interesting extension would be redoing the analysis and the model building process with a larger and more comprehensive dataset. Incorporating additional baseline variables, such as detailed clinical data, pre-existing medical conditions, and blood oxygenation levels, may significantly enhance the predictive power and the interpretation of the model.

## Data Availability

All data produced are available online at https://data.mendeley.com/drafts/7bmfgkkm3z

## 7 Appendix

All of the R code and dataset used for this paper can be found in https://github.com/ZhilongZ2/MS-Thesis-code.

## 8 Supplementary Material

This section contains additional figures referenced in the main text. These supplementary materials provide further insights and support the findings discussed in this paper.

